# A prevaccination validated network that drives the breadth of the protective neutralizing antibody response following Dengue Vaccine TV003 immunization

**DOI:** 10.1101/2021.09.25.21264123

**Authors:** Adam Nicolas Pelletier, Gabriela Sanchez, Mark Watson, Abdullah Izmirly, Tiziana Di Pucchio, Elias Haddad, Eustache Paramithiotis, Jorge Khalil, Michael Diamond, Esper Kallas, Rafick Pierre Sekaly

## Abstract

Development of fully protective dengue virus (DV) vaccines has been problematic as infection with DV requires a broad antibody immune response that targets all 4 possible serotypes. Herein, we used an integrated systems vaccinology approach to identify prevaccination features that allow the development of fully protective DV-specific antibody responses. This approach allowed us to identify a transcription network in a subset of monocytes defined by the expression of CD68 and downstream of specific pro- and anti-inflammatory cytokines. Moreover, we identified metabolites as drivers of an immune response that induced neutralizing antibodies to the 4 DV serotypes. Specifically, PC/PE drove the production of TGF-B in CD68 ^low^ monocytes, which was a positive correlate of the protective antibody response. In contrast, primary and secondary bile acids triggered a proinflammatory response downstream of TGR5 signaling and inflammasome activation in CD68 high monocytes, which was associated to a non-protective antibody response. These features were validated in vitro in primary myeloid cells. Our results highlight the role of cell and systemic metabolism as regulators of protective immune responses to vaccination, and that systems vaccinology is a key tool to identify such mechanisms.

## INTRODUCTION

Dengue infectious disease is a mosquito-borne viral disease caused by one of the four serotypes of Dengue viruses (DENV-1, 2, 3 and 4). At present, this disease is endemic in more than 100 countries, mainly in tropical and subtropical regions, with the highest incidence of infection in southeast Asia, South and Central America (WHO). Clinical manifestations of Dengue infection can range from an asymptomatic or mild disease to dengue hemorrhagic fever (DHF), a life-threatening syndrome associated with multi-organ failure. Although any DENV serotype can cause severe disease with a primary infection, a secondary infection with a heterologous DENV serotype has been associated with an increased risk of DHF. This phenomenon has been linked to Antibody-dependent enhancement (ADE), in which primary antibodies facilitate the secondary infection by increasing the uptake of non-neutralized DV virions by monocytes or macrophages via the Fc-gamma receptor.

The first authorized vaccine against DV, Sanofi Pasteur’s tetravalent live attenuated Dengvaxia (CYD-TDV) was approved in Dengue-endemic countries throughout Asia and Central/South America in 2016, with a vaccine efficacy differing by serotype, age group. However, the largest benefits were found in children with a positive prevaccination serostatus, whereas vaccinated seronegative children (<9 yrs old) were found to have an increased likelihood of hospitalization for severe dengue. This has since been suggested to be caused by ADE in a way akin to that of a primary infection: where a neutralizing antibody response against only a subset of all 4 serotypes is a risk factor for severe dengue.

There is an ever increasing body of evidence investigating the heterogeneity of immune responses induced by those vaccine approaches across individuals, both in regards to its quality and magnitude. The so-called “immune predisposition” to vaccination has been associated to immune subset frequency and distribution and circulating levels of cytokines and chemokines both at prevaccination and early post-vaccination timepoints. Moreover, these features are also known to be further modulated by a combination of age, gender, genetics and a complex interplay between the microbiome and cell metabolism, in order to ultimately influence global health.

Systems vaccinology approaches have provided valuable insights into the identification of correlates of immunogenicity and protection, by taking advantage of the wealth of data from large “OMICs”, including transcriptomics, proteomics and metabolomics, in combination with unbiased predictive computational analyses. This has led to the successful identification of a predictive early gene signature of the antibody response in the yellow fever live-attenuated vaccine YF-17D, influenza vaccine and HepB vaccine (. Moreover, systems vaccinology has been leveraged to isolate pre-vaccination signatures in the context of Influenza, HepB, Malaria and Yellow Fever vaccination.

We leveraged multi-omic data from a Phase II clinical trial for the tetravalent live-attenuated TV003 vaccine (NIAID/NIH) in Brazil to identify correlates of the breadth of the neutralizing antibody (nAb) response. A combination of transcriptomic, metabolomics, proteomics, flow cytometry and in vitro validation identified a complex immunometabolic balance between pro- and anti-inflammatory signatures mediated by monocytes predisposing to an optimal response in seronegative participants.

## RESULTS

### Breadth as a feature of optimal vaccine response in the TV003 Phase II clinical trial

The induction of a broad nAb response to all 4 DENV serotypes is required to be the best vaccination strategy to prevent ADE in seronegative individuals who are at risk of a secondary infection. The vaccination strategy of the TV003 Phase II clinical trial is detailed in Supplementary Figure 1A. A total of 46 seronegative individuals were enrolled in the trial and received two rounds of immunization with the second dose given 180 days after the first dose. Supplementary Figure 1B shows that the vaccine was shown to induce a potent multivalent response in the majority of individuals for at least 90 days. Moreover, we observed this response to be mostly restricted to the primary immunization, as we did not witness a secondary increase in nAb titers post boost. The kinetics and magnitude of this primary response varied significantly across serotypes and participants (Supplementary Table 1). We identified 3 non-responders (8.51%), whose serum could not neutralize any of the 4 serotypes in PRNT assays at any timepoint before Day 90. In responders, we observed the peak response, defined as the earliest maximum titer timepoint per participant, to be at Day 28 for a close to half of participants for DV1 and DV3 (48.4% and 54.8% respectively), while the remainder are spread between Days 56 or Day 91. In contrast, the distribution of titer peaks for DV2 and DV4 displayed an enrichment for Day 56 maxima (45.2% and 48.4% respectively), despite also showing substantial heterogeneity. To determine if this was driven by a patient effect, namely if some participants responded later for all serotypes than others who peaked earlier, we tallied the number of peak timepoints per participant (Supplementary Table 2): we highlight that at least half of all responders (54.5%) have 2 timepoints at which they have their personal maxima and 33% display peaks at all 3 timepoints. Only 4 participants (12.1%) exhibited a simultaneous response across serotypes, suggesting this heterogeneity in nAb kinetics is not caused by a patient effect, but by unexplained differences across serotypes.

In order to integrate this information and circumvent this large heterogeneity and infer a measure of the magnitude of the response per participant, we measured the log-transformed Area Under their nAb titer Curve (AUC) using the trapezoid rule. In parallel, we calculated our selected outcome, namely the Breadth of the nAb response, in Supplementary Table 3. In summary, breadth was calculated on a binary basis of detection / absence (nAb >10) for each serotype for at least one timepoint (columns 2-5), and then summed up across serotypes, as per the clinical definition(column 6). The outcome was then dichotomized into high and low breadth groups, where high corresponds to a detectable nAb response to all 4 serotypes, and low to values under 4 (column 7). This choice was made on the basis of the biology of ADE: anything below a full breadth brings a potential negative risk for ADE, and was thus considered suboptimal.

A majority of participants (26/35, 74.2%) had successfully mounted a high breadth response at Day 90 (Suppl Fig 1C), with the remaining with either a partial (6/35, 17.14%) or absence of response (3/35, 8.57%),

Examination of the low breadth responses showed that all participants retained the capacity to mount a detectable nAb response against DV1, suggesting that the immunization for this serotype was the most consistent and robust. In contrast, DV3 responses seemed to be particularly lacking in low breadth individuals, where 7/9 participants (including 3 non-responders to all 4 serotypes) displayed no detectable nAb titers against DV3. Moreover, comparison of the magnitude of the response across breadth groups showed a partial association between outcomes (Supplementary Figure 1D), where DV3 AUC was significantly lower in the low breadth group (p = 0.00023), along with a weaker significant association of DV2 and DV4 AUC to Breadth (p = 0.16, p = 0.022, respectively). This supports that the induction of a DV3-specific response is a critical determinant of Breadth following TV003 immunization.

### Type I Interferon and TGF-B signaling dichotomy as inflammatory drivers of breadth of the nAb response to TV003

Next, we performed a bulk RNAseq analysis to evaluate the transcription profile of prevaccination PBMC (Figure 1A). Unsupervised Principal Component Analysis (PCA) reveals no significant association of the top components with the AUC and Breadth outcomes (data not shown). However, using a supervised analysis (DESeq2), we detected a total of 1139 differentially expressed genes (DEGs) at Day 0 associated with the breadth of the nAb response before vaccination (nominal p-value < 0.05) (Figure 1A), where 530 and 609 were significantly associated with a high and low breadth, respectively.

**Figure 1.**
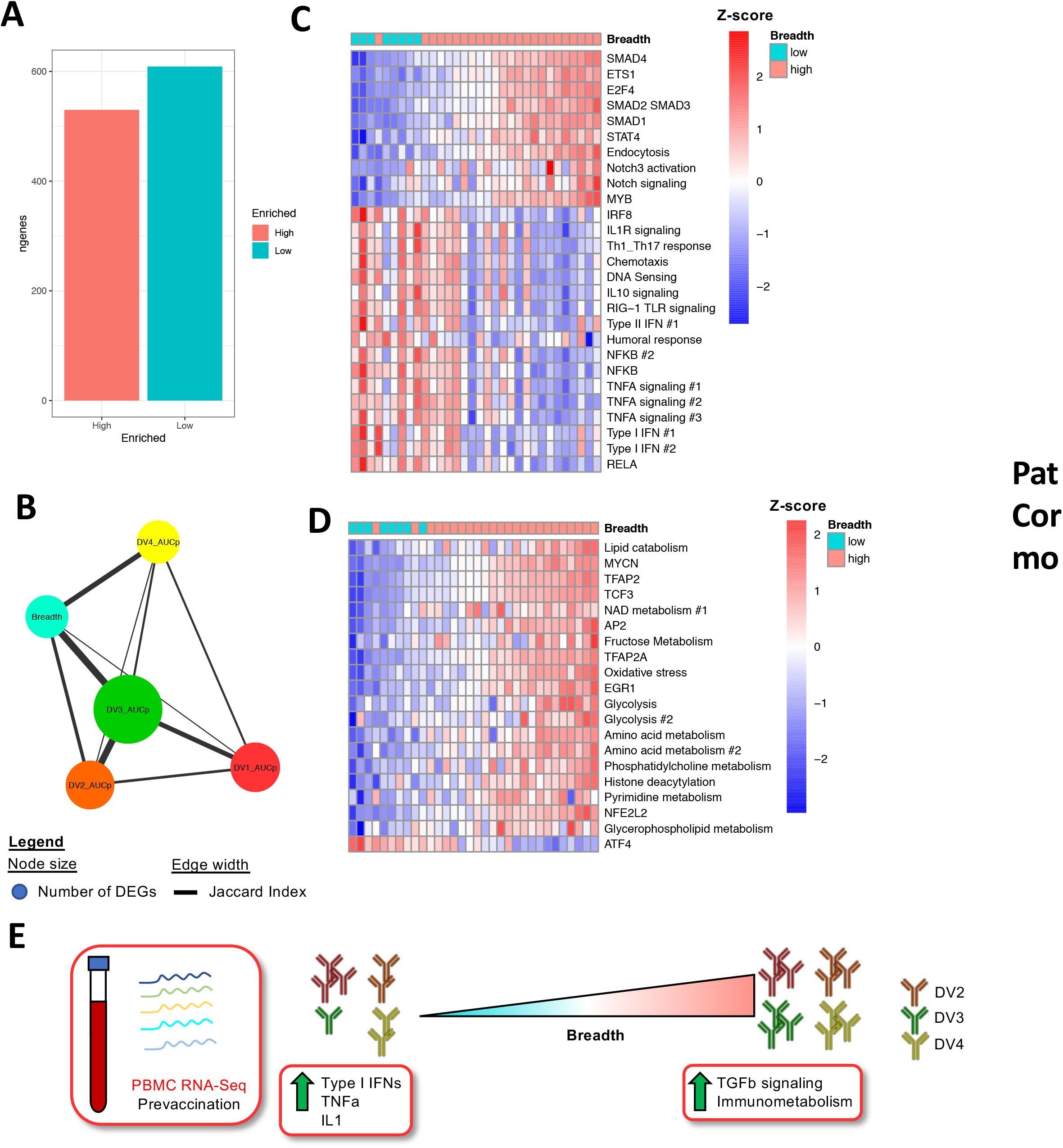
PBMC transcriptomic analysis of prevaccination of TV-003 vaccine-induced Breadth of the response reveals a dichotomic association of pro vs anti-inflammatory pathways. (A) Heatmap representing row normalized 1139 DEGs associated with Breadth of response at a nominal p.value <= 0.05, where each row represents a gene and columns represent prevaccination timepoint samples. (B) Jaccard similarity network across outcomes for transcriptomic contrasts. Each node represents an outcome, such as Breadth of the response or the magnitude of serotype-specific responses, and edge widths represent the Jaccard index of DEGs found to be significantly associated at p.value < 0.05 in each pairwise comparison. Heatmaps representing pathway EnrichmentMap analysis for immune (C) and metabolic (D) transcriptomic genesets, where each row represents a pathway module and rows individual participants. Row annotation tracks (left) represent the signed log p-value of the association of each outcome for each module, with blue and red denoting high negative and positive significance, respectively. Column annotation tracks (above) represent the Breadth and DV3-specific AUC outcomes per participant.

Parallel DEG analyses for each serotype-specific outcomes revealed more insight into the relationship between those outcomes (Figure 1B). The number of DEGs differed drastically across outcomes, identifying 1471, 1434, 2427 and 1155 DEGs respectively for DV1-4, with DV3 displaying the highest number of DEGs. Examination of the DEG similarity across outcomes, using the Jaccard coefficient, highlights the high DEG overlap between DV3 and Breadth (465), and low overlap of DV1 with Breadth (94).

We then performed a pathway analysis using pre-ranked GSEA, which revealed significant associations of pathways with outcome, as quantified by a NES (Normalized Enrichment Score). We identified a total of 3489 genesets significantly associated at a nominal p-value < 0.05 (2849 associated with high breadth, 625 with low breadth), or 1039 at an adjusted p-value (FDR) < 0.05 (799 high; 240 low). EnrichmentMap was used to generate modules on the basis on the geneset leading edge Jaccard similarity index cutoff of 0.25. We identified 27 immune modules (Figure 1C) 10 of which were associated with a high breadth and 17 with a low breadth, along with 20 metabolic modules (Figure 1D) (19 high; 1 low). Our analysis revealed a positive association of several anti-inflammatory genesets associated with TGFB signaling, namely SMAD2/3/4 and ETS1, with p-values ranging from 0.008 to 0.0002. In contrast, we identified a negative association of several proinflammatory genesets with Breadth, with the strongest association being Type I interferons, TNFA and RELA modules (p < 0.01 - 0.00018). Examination of the leading edge genes for those signatures highlights a predominance for antiviral function (IFIH1, DDX58, IRF1,IFIT1, IFIT2, IRF7) and tonic signaling ISGs (STAT1, USP18, PARP9) (Supplementary Fig. 4).

Examination of gene expression levels for cytokine and chemokine expression did not however reveal a significant association of TNFA, type I IFNs or TGFB at a univariate level in relation to the Breadth outcome, nor are they found within the leading edge genes of modules generated in Fig.1C. This suggests either the source for these cytokines is either not blood-borne/hematopoietic, comes from a subset too rare to pick up the single signal from or that the effect is being diluted by the subset heterogeneity. In contrast, a few cytokine genes highlighted in our pathway analysis also proved significant at a univariate level, including IFNG and multiple members of the IL1 family (IL1B, IL1A, IL6). This suggests a differential activation of a member of the inflammasome family between high and low breadth to be investigated. Finally, we identified 10 chemokines enriched in the low breadth group and 1 in the high breadth group, which was consistent with the general Chemotaxis module being negatively associated with Breadth in Fig.1C.

Given the large known interplay of cell metabolism and immune function and the balance of pro vs anti-inflammatory pathways highlighted above, we investigated whether prevaccination transcriptomic measurements of metabolism could also predict post-vaccination Breadth (Fig. 1D). We identified a large number of very biologically diverse pathways significantly associated with Breadth, including NAD metabolism, Oxidative Stress, Glycolysis and Amino Acid metabolism as positive determinants of a good response. The highest associations were however for modules involved in lipid metabolism, including transcription factors such as TFAP2 and TCF3 (p-values ranging from 0.0006 - 0.00033).

We measured the concentration levels for a custom panel of 228 plasma protein using monitoring-mass spectrometry (MRM) analysis. Notably, we identified 13 plasma proteins to be significantly associated to the breadth of the response at a univariate level (p < 0.1), 10 of which were associated with a low breadth of nAb responses (Figure 2B). These proteins were hallmark surrogates of proinflammation, including Complement receptor 1 (CR1; p=0.0066), soluble ICOSL (p=0.028), C-Reactive Protein (CRP; p=0.08), Lysozyme C (LYSC; p=0.028) and Complement Factor 1 (CFAI; p = 0.026). Furthermore, know immunometabolic determinants were also identified in the plasma, such as FABP5 (p = 0.09), NGAL (p = 0.007) and Serum Amyloid 1(SAA1; p=0.076), all of which are involved in lipid metabolism.

**Figure 2.**
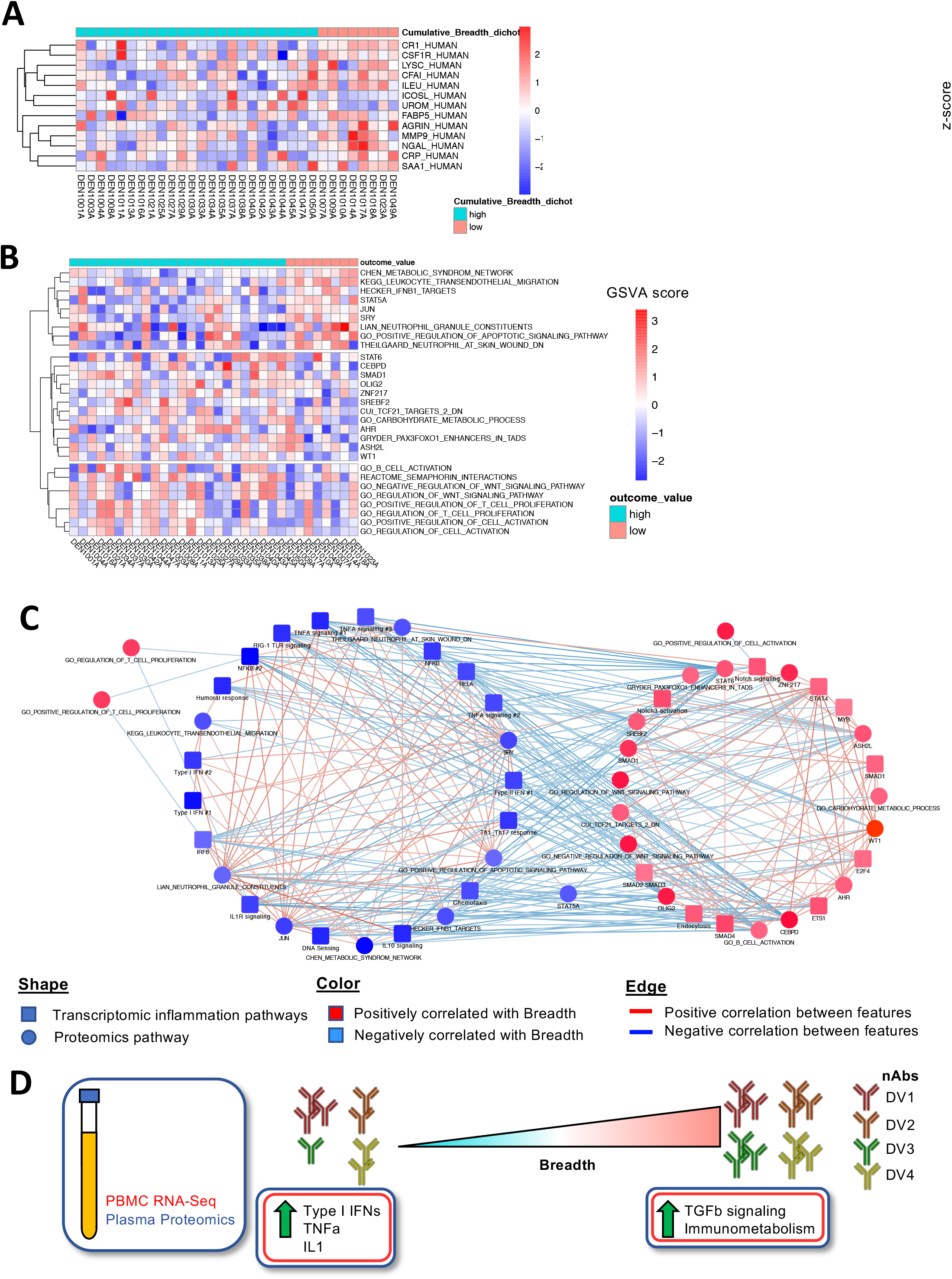
Prevaccination plasma protein profiling confirms a negative association of proinflammatory pathway signaling at the protein level. (A) Heatmap representing Mesoscale quantification of row-normalized prevaccination plasma chemokines and cytokines associated with Breadth of the response to TV-003 at a nominal p.value <= 0.05 (left) with associated boxplots per significant analyte (right). (B) Differentially expressed plasma proteins between high and low Breadth patients by MRM proteomics at p.value < 0.05. Row represent individual proteins and their UniProt accession ID. The color scale, from blue to red, represents the normalized expression (z-score) of the associated row, where red denotes high expression. (C) Geneset Variant Analysis (GSVA) on MRM proteomics data heatmap. Rows represent the GSVA score per participant for the combination of the proteins in the enriched pathway. Pathways were enriched at a nominal p-value of 0.05 (D) Integrated correlation network analysis of transcriptomics and proteomics, where nodes denote features and edges the Spearman correlation coefficient between them.

Further investigation of the proteomic signature using Gene Set Variation Analysis (GSVA) on all proteins revealed a number of differentially enriched pathways associated with breadth, among which we isolated HECKER IFNB1 targets (p=0.026) and SMAD1 (p=0.0156) pathways (Figure 2C), which respectively served as protein validation of the dichotomy of IFN and TGFB signatures identified by RNA-Seq in Figure 1C. Moreover, we noted an interesting enrichment of plasma proteins involved in B cell activation (p=0.04) in high breadth participants. Integrative analysis between transcriptomics and proteomics supports a strong consistency across OMICs and highlights a consistent dichotomy between pro and anti-inflammatory in association with the breadth of the response to TV003 (Figure 2C).

### Pre-vaccination metabolic activity as immunomodulating driver of vaccine response to TV003

Given the growing recognition of cell metabolism as modulator of immune responses following vaccination and our transcriptomic observations for an association of metabolic activity with breadth of the response, we hypothesized that cell metabolism was a critical driving factor for the divergent pro- and anti-inflammatory profiles identified in Figure 1 prior to TV003 vaccination. Given the large breadth of metabolic pathways known to possess immunomodulatory function, we analyzed a broad panel of 868 pre-vaccination plasma metabolites in relation to the breadth response (Supplementary Figure 5A). Despite substantial heterogeneity across participants, which is a common feature of both plasma metabolism and baseline donor characterization, we identified 22 differentially expressed metabolites (nominal p-value <0.05) across breadth groups, 6 of which belonged to the Phosphatidylcholine/Phosphatidylethanolamine (PC/PE) class of metabolites. Interestingly, PC/PE has been previously associated to TGF-B function, both by regulating its transcription and by contributing to its activation at the cell surface in chondrocytes.

We then proceeded to perform pathway analysis using MSEA (Metabolite Set Enrichment Analysis) on SMPDB (Small Molecular Protein DataBase) and Metabolon metabolite sets, which foregoes traditional p-value thresholds and operates by ranking the whole list of metabolites for enrichment, followed by Enrichment Map (as described above for transcriptomics) for consolidating of sets with high degrees of redundancy. We further showed the association of PC/PE metabolism as a positive correlate of the breadth response (p = 0.00037) (Figure 3B), in line with the univariate observations described previously. We highlighted the strongest drivers of module enrichment from the leading edge of correlates in Figure 3A; PC/PE metabolism includes 14 metabolites. Other positive module correlates included Benzoate and sphingomyelin metabolism (p=0.005 and p=0.013). In contrast, we observed a significant association of bile acid metabolism with breadth of the response, with primary Primary Bile Acids cholate and glycocholate as the most potent features, as negative correlates of the response (p=0.00049).

**Figure 3.**
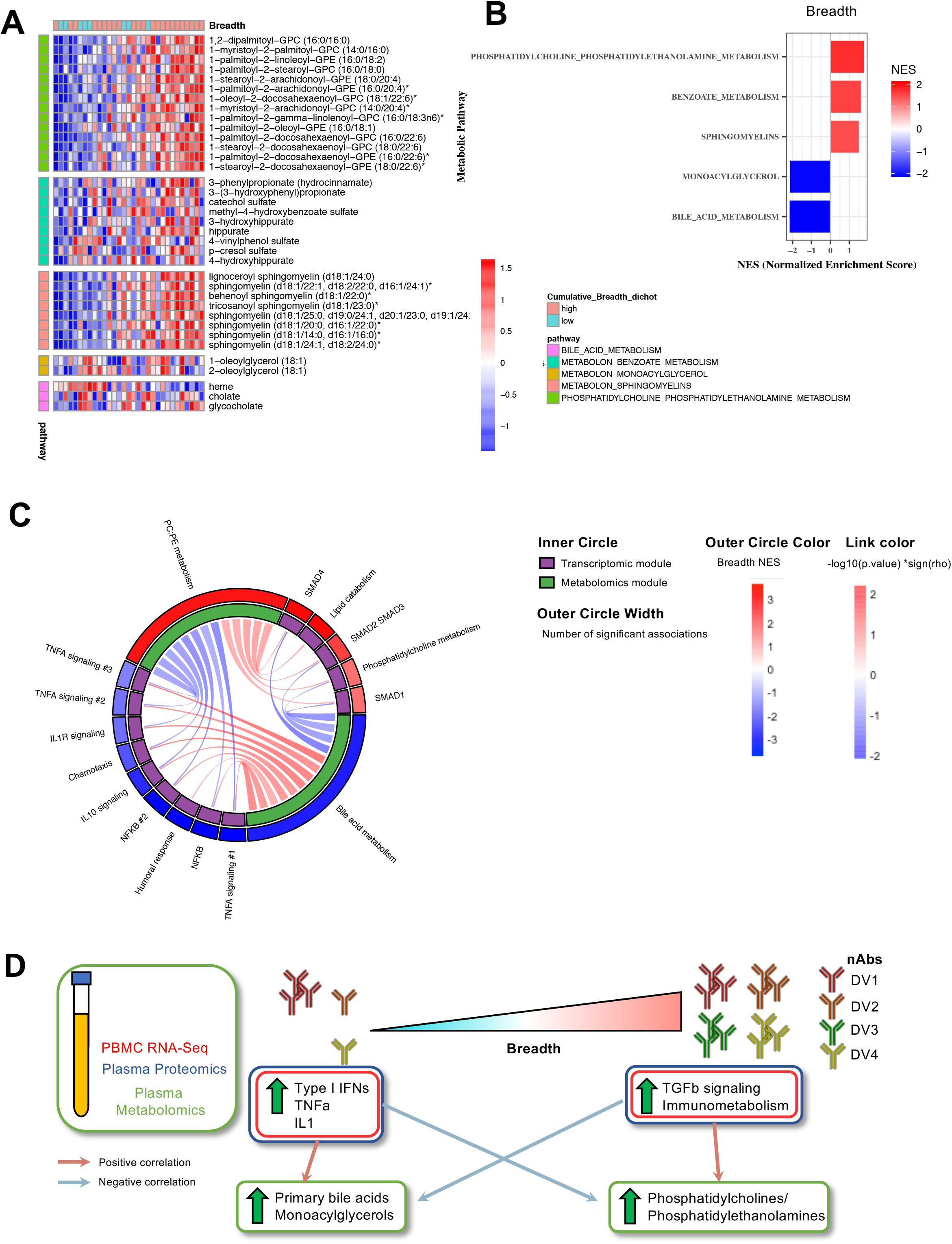
Prevaccination profilling of plasma metabolites reveals metabolic pathway activity is associated with inflammatory status. (A) Metabolite Set Enrichment Analysis (MSEA) was performed on preranked metabolites from differential expression analysis, followed by Enrichment Map analysis with a Jaccard Index > 0.5 for modules displaying a nominal p.value < 0.05. (B) Heatmap representing the leading Edge metabolites from modules in A, where each row represents normalized metabolite expression and columns individual participants. Row annotation tracks (left) represent the signed log p-value of the association of each outcome for each module, with blue and red denoting high negative and positive significance, respectively. Column annotation tracks (above) represent the Breadth and DV3-specific AUC outcomes per participant. (C) Integrated correlation network analysis of transcriptomics and metabolomics, where nodes denote features and edges the correlation coefficient between the 2 features they link. Spearman correlation (p-value < 0.05) was used as an integration metric across OMICs and displayed on the edge color. Node shape and color maps to the respective feature type and association to Breadth.

We performed integrative analysis between transcriptomic and metabolic sets allowed to confirm the concordance between metabolic pathways across OMICs, where PC metabolism had been highlighted in RNA-Seq (Figure 1C) as a predictor of breadth of vaccine induced Ab responses (Figure 3C). Spearman regression shows a significant correlation between PC/PE metabolism pathways across OMICs, validating our prior observations. Furthermore, this analysis highlighted the interplay between metabolic pathways and cytokine activity, where PC/PE metabolism and Bile acid metabolism modules are respectively negatively and positively correlated to 13 proinflammatory modules, including TNFA signaling, ILR1 signaling and NFKB. On the other hand, those metabolic modules are also correlated to 5 positive transcriptomic determinants of breadth, 3 of which are involved in TGFB signaling (SMAD1, SMAD2/3, SMAD4). These observations serve to further display the immunometabolic dichotomy associated with an optimal response to vaccine TV003.

In order to pinpoint the potential immune populations that could be mediating bile acid signals and PC/PE activity, we leveraged publicly available sorted subsets from the Monaco dataset (Supplementary Figure 6). We infer myeloid cells, with monocytes in particular, as the most probable subset involved in bile acid metabolism, on the basis of their predominant expression of the canonical TGR5 (Takeda G-proOtein-Coupled Receptor 5 protein, encoded by GPBAR1) receptor and of the VDR. Moreover, despite FXR’s well-established role in mediating downstream signaling of bile acids, we could not detect its expression in any of the immune populations across datasets. In contrast, PE/PC metabolism has no established receptor, and is thus more difficult to associate to a specific subset in a bulk RNA-seq dataset. In order to pinpoint the most likely mediators of PC/PE metabolism in the blood, we generated an aggregate score from the normalized expression for each sorted subset in the Monaco dataset. This allowed us to observe an enrichment for both PC/PE metabolism and bile acid metabolism in the myeloid compartment, notably in monocytes. We also observed an elevated expression of genes in both of these pathways in progenitors and plasmablasts. We were intrigued in the lower relative expression of the bile acid pathway genes of Monocytes and mDCs, in comparison with Progenitors and Plasmablasts given the expression of the canonical TGR5 receptor in Supplementary Figure 6A. While we cannot exclude a plasmablast contribution in the response, the inflammatory signatures identified previously are generally associated with innate immunity, and as such makes the myeloid compartment more likely mediators of the prevaccination predisposition effect we observe.

### Prevaccination classical monocyte functional heterogeneity is associated to breadth of response to TV003

In light of the signatures identified previously, we hypothesized a major contribution of the innate compartment in the predisposition of an optimal response to TV003. We set out to associate prevaccination PBMC flow cytometry profiles of innate immune cells to the breadth of Ab responses. Unsupervised clustering using a combination of Phenograph clustering with tSNE dimension reduction approach revealed 22 distinct clusters amongst CD3^-^CD19^-^CD56^-^ populations (Figure 4A). The frequencies of 3 of these clusters were significantly associated with the Breadth of the response: one of which was consistent with a pDC phenotype (Lin^-^ CD11c^+^CD123^+^; cluster 8, p = 0.0062) and the other 2 clusters with a classical monocyte phenotype (CD14^+^CD16^-^CD11b^+^HLA-DR^+^) (Figure 4B; Supplementary Fig.7A)). However, those 2 monocyte clusters, which differed primarily by their expression of scavenger receptor CD68 (macrosialin), and with a lesser extent by CD45 expression, displayed a differential enrichment across breadth groups in the DV trial. CD68^hi^ monocytes (cluster 1) were enriched in the low breadth group (p=0.0028), whereas CD68^low^ monocytes (cluster 16) were enriched in the high breadth group (p = 0.0024).

**Figure 4.**
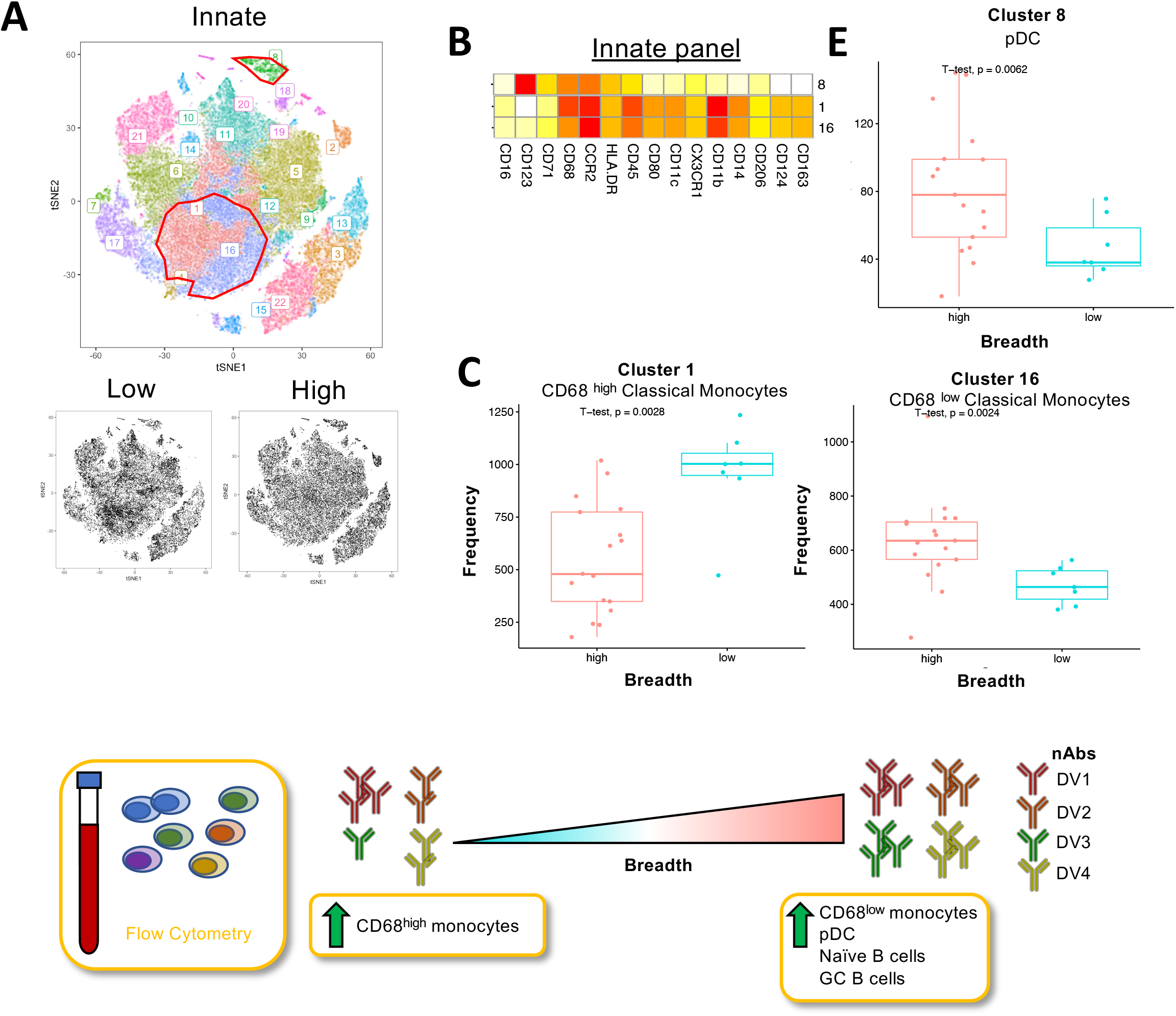
The frequency of prevaccination classical monocyte subsets distinguished by their expression of scavenger receptor CD68 at the prevaccination timepoint is associated with Breadth of the response to TV-003 in a dichotomous fashion. (A) tSNE plot of CD3-CD19-CD56- PBMCs from unsupervised flow cytometry analysis. Different colors represent distinct clusters following Rphenograph clustering analysis (top). The tSNE plot was separated on the basis of breadth outcome (bottom), showing variations in density in clusters 1, 16 and 8 (surrounded top). (B) Heatmap showing scaled median fluorescence cluster profile for each marker: each row is a significant cluster and each column is a marker in the panel. White denotes low expression, while red denotes high expression. Red rectangle highlights the top 2 significant correlates of breadth (C) and (D) Boxplots and scatter plots, respectively showing cluster 1(top) and 16 (bottom) cell frequency versus Breadth and DV3-specific AUC. (E) Boxplot showing cluster 8 cell frequency versus Breadth.

Furthermore, flow cytometry characterization of the B cell compartment highlighted additional differences between high and low breadth participants. Our B cell panel lead to the identification of 27 CD19^+^ clusters (Figure 4E), 2 of which were significantly enriched in the high breadth group. Cluster 8 was consistent with a Naive B cell phenotype (CD20^+^CD21^+^BCL2^hi^IgG-), while Cluster 15 was consistent with a GC B cell phenotype (CD20^hi^CD21^low^CD38^+^) (Figure 4F; Supplementary Fig 7B).

In order to determine if prevaccination CD68 expression in CD14+CD16-monocytes was associated with other vaccine responses, we leveraged CITE-Seq data from Tsang et al. generated in the context of influenza vaccination across 20. subjects (Figure 5A). Subjects that showed the highest level of CD68 expression also had the highest magnitude off the antibody response (p = 0.03)(Figure 5B), confirming the association of CD68 expression and protective responses to DV vaccination. MAST analysis identified multiple genes (2160 with FDR <0.05) associated with CD68 expression amongst CD14+CD16-monocytes. We observed a positive association to expression of bile acid receptor TGR5 (GPBAR1), suggesting CD68^high^ monocytes could be more responsive to bile acids. Furthermore, we associate a number of proinflammatory genes to CD68 expression, including TNF, MYD88, and PYCARD (inflammasome activator ASC). Moreover, we showed that members of the antiviral interferon network (Suppl. Fig.4) IFIT1/2 and IRF1 were positively associated to CD68 expression. Conversely, we identified a negative association of anti-inflammatory myeloid scavenger receptor CD163 to CD68 expression. These observations highlight the impact of differences in CD68 expression in monocytes, a significant correlate of DV-specific protective antibody responses on mechanisms that underlie the development of those antibody responses.

**Figure 5.**
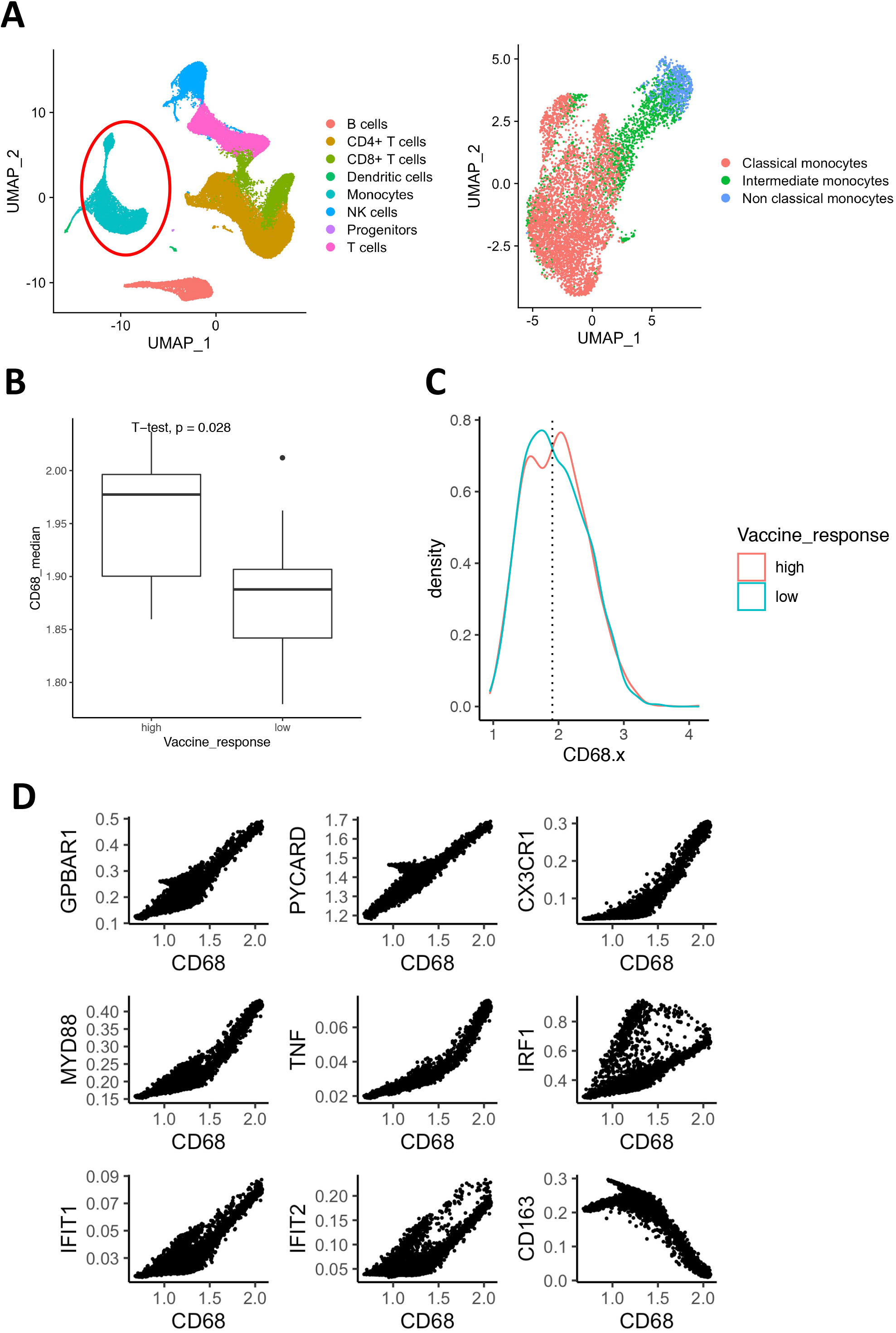
Single-cell transcriptomics analysis on 4 independent datasets confirm existence of novel subsets of monocytes with distinct proinflammatory vs anti-inflammatory functions. (A) CD68 gene expression violin plots across monocyte clusters, where each dot represents a cell in the associated cluster. (B) GSEA analysis of between CD68^low^ Classical monocyte cluster 3 versus CD68^high^ Classical monocyte clusters 1, 2, 4 and 5, where a positive NES (red) indicates enrichment of genes in associated pathway in Classical Monocytes #2, and NES <0 (blue) in Classical Monocytes #1 (C) Heatmap of row-normalized leading edge genes from each enriched pathway across selected clusters for Type I IFN (top) and SMAD (bottom) pathways.

**Figure 6.**
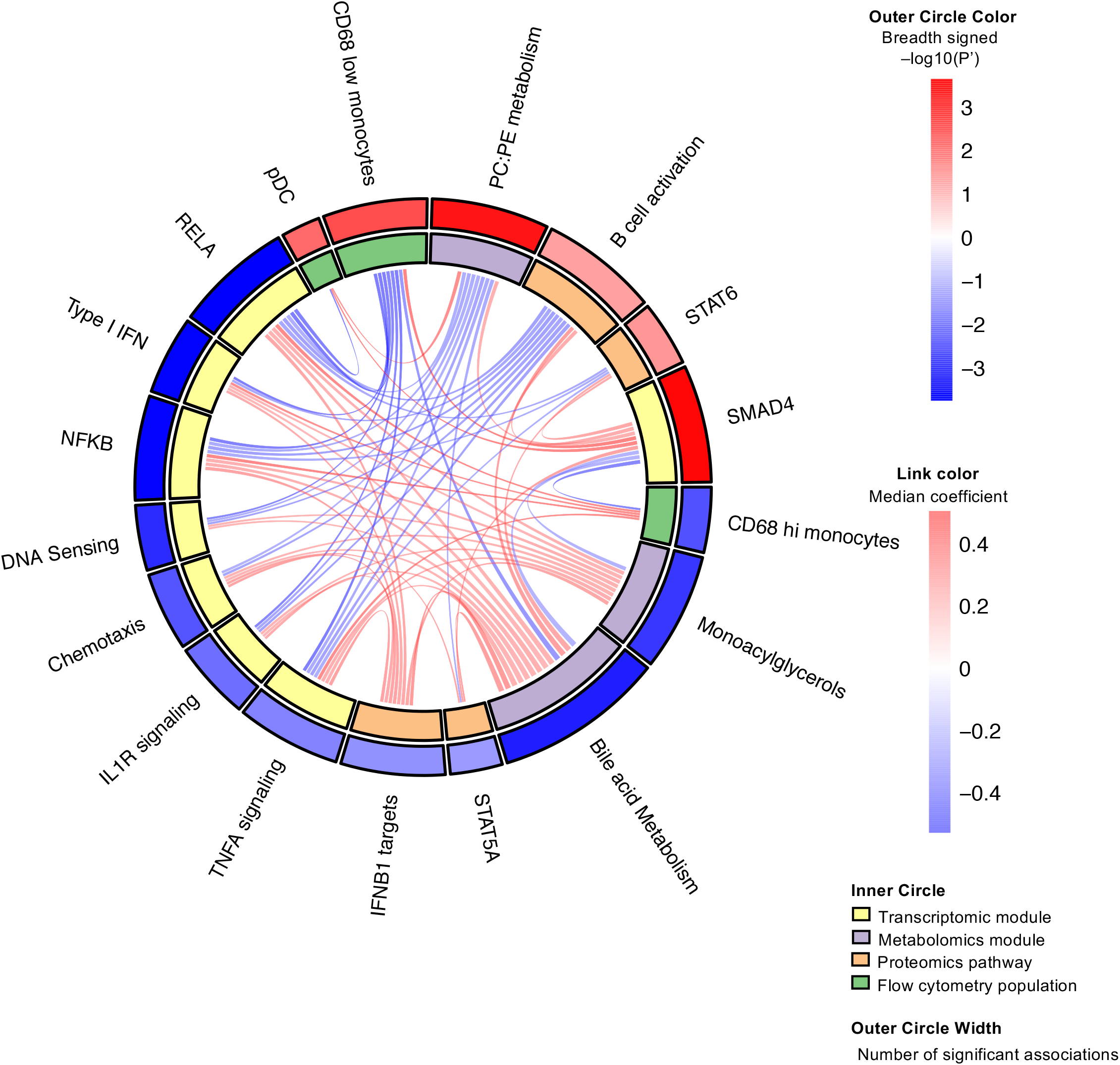
Integrative analysis between transcriptomics, Plasma Proteomics and Metabolomics and flow cytometry highlight a dichotomous crosstalk of classical monocyte subsets with metabolic, pro and anti-inflammatory pathways. Circos plot representation of cross-platform correlation analysis, where nodes denote features and edges the correlation coefficient between the 2 features they link.

### Integrative analysis of TV003 OMICs reveals monocytes as central mediators of an inflammation-metabolic crosstalk

We used sparse least-square regression models to investigate whether the gene expression signatures associated to breadth in the TV003 Phase II trial were correlated to levels of plasma proteins, plasma metabolites and FCM subset frequencies, both at the single analyte or pathway level (Supplementary Figure 9 and Figure 7, respectively). The pathway-level integrative analysis shows a clear dichotomy between pro- and anti-inflammatory pathways and their respective association to CD68^high^ and CD68^low^ classical monocytes. For instance, they are respectively negatively and positively correlated to the SMAD4 module, hinting of an enhanced TGFB signaling in CD68^low^ monocytes. pDC are also positively associated to SMAD4 and not correlated to Type I IFN, providing some potential mechanistic insight into a more tolerogenic pDC phenotype as a positive determinant of breadth. Interestingly, the frequency of neither monocyte population are correlated to the metabolite modules, yet are connected to the same functional inflammatory modules (SMAD4; Type I IFNs, RELA/NFKB, TNFA signaling).

## DISCUSSION

In this study, we leverage an in-depth multi-OMIC analysis to provide a mechanistic framework underlying the predisposition to an optimal response to a live attenuated tetravalent vaccine to dengue virus. This Phase II trial included 38 seronegative participants that were vaccinated with the TV003 vaccine in Brazil. We identified an intricate network of bile acids and PC/PE metabolites respectively triggering activation of pro- and anti-inflammatory signaling cascades in monocytes. This in turns modulates how many DV serotypes are neutralized post-vaccination. i.e. Breadth of the response.

Associations of the anti-inflammatory properties of PC/PE have been made in the past by showing their role in promoting downregulation of TNFa signaling and autophagy. PE deficiencies have been associated to the Unfolded Protein Response (UPR), ER stress and in the context of Parkinson’s disease and ulcerative colitis. The mechanisms through which PC/PE induce these anti-inflammatory effects are however poorly defined. One proposed mechanism highlighted through in vitro rat chondrocyte experiments has been through activation of latent TGF-beta1 in a manner similar to mild detergents. Our validation experiments have been able to further validate these findings with human primary monocytes, showing both an increase in active TGFb concentrations in supernatants following exogeneous PC/PE exposure and SMAD2/3 phosphorylation downstream of TFBGR signaling. Interestingly, a recent report by Fu et al. highlights a role for PE in promoting CXCR5 expression and differentiation of T_FH_ cells. This could be in fact mediated through TGFb as well, which is a well known key factor in T_FH_ differentiation. This could also mean PC/PE play a role beyond monocytes in the context of our cohort, optimizing T_FH_ differentiation and facilitating the generation of a broad nAb response.

In contrast, exogenous bile acid stimulation was associated to a pro-inflammatory phenotype in human primary monocytes. We detected downstream signaling of bile acid receptor TGR5 by CREB1 phosphorylation, as well as inflammasome activation through Caspase-1 phosphorylation. We find this interesting because, despite having Interferon and TNFA as more significant pro-inflammatory transcriptomic pathways in association to Breadth, we do not detect any differential expression of the genes encoding for the cytokines in these pathways, thereby suggesting their source is either non-hematopoietic or from a rare subset within PBMC. In contrast, almost all IL1-related cytokine genes were found to be enriched in low breadth participants PBMC, which better support a hematopoietic source of this inflammasome activity. The role of bile acids in inducing an inflammatory response has a complicated history: work in mice has shown FXR and TGR5-mediated mediated inhibition of NLRP3 activation downstream of cAMP-PKA-CREB in bone marrow-derived macrophages, as well as a tendency to promote a M2-like phenotype with increased IL-10 production for both primary and secondary bile acids. Furthermore, the discrepancy with our results may be mediated by the species difference: authors have shown that their inhibition of NLRP3 requires activation of intracellular bile acid receptor FXR, for which we detect no transcripts in human monocytes in the Monaco and Human Cell Atlas datasets. Furthermore, additional conflicting evidence also purports a more proinflammatory role of bile acids, including an activation of the PYRIN inflammasome in human PBMC by bile acid analog BAA473, leading to increased IL-18 and IL-1b secretion and activation. Bile acids have also been shown to promote hepatocyte cell death, release of alarmins in cholestasis patients, a condition associated with an accumulation of bile acids in the liver. Elevated expression of inflammasome genes was observed in both hepatocytes and M1-like macrophages in cholestatic patients, supporting our observations of a pro-inflammatory role of bile acids.

The role of bile acids in the context of vaccine responses has been described previously. Hagan et al. have shown that gut microbiome perturbations alter immunity to vaccines, partially through modulation of secondary bile acids in the context of influenza vaccination (TIV). In this study, bile acids were not directly associated to post-vaccination outcome, but were predominantly negatively associated to inflammasome signaling. This finding comes in contrast with ours, which could be explained by gut microbiome disparities due to geographical location (California versus Brazil), and the distinct nature of the vaccine. As a matter of fact, this study contrasts with other published and unpublished work that highlight a different role of inflammatory signatures across vaccine regimens. To support this statement, a recent study by Liu et al. uncovered hypo-responsiveness to HBV vaccination in children with elevated bile acids, which is in line with our findings.

The dichotomy of pro- and anti-inflammatory signatures as respective negative correlates of vaccine immunogenicity has been the focus of much of the work published in the context of vaccine predisposition. A Study by Fourati et al. has demonstrated hyporesponsiveness to HepB in the presence of heghtened inflammation. On the other hand, work by the HIPC, currently under review, has shown a pan-vaccine proinflammatory signature as a positive correlate of the magnitude of the Ab response. We believe the differences lie in the necessary response for each vaccine. For instance, an individual displaying an enhanced level of immune activation, expression of antivirals would also limit viral replication of a live attenuated vaccine like TV003, effectively reducing antigen load for the generation of a potent humoral response. Moreover, we think that the term proinflammatory response may be too reductive: a heightened NFKB differ greatly in downstream pathways induced to that of RIG-I/MDA5 signaling.

## MATERIAL AND METHODS

### Trial design

Thirty-seven volunteers from TV003 Phase 2 trial in Brazil were used in this study, based on their prevaccination seronegative status to all 4 DV serotypes as measured by PRNT assay by the Kallas laboratory in Sao Paulo. Participants were recruited at the Butantan Institute in Sao Paulo, were screened for major infections prior to vaccination, and were given 2 doses of TC003 vaccine spaced 180 days apart. PBMCs and plasma were collected at Day 0, prior to vaccination, as depicted at Supplementary Figure 1A. Neutralizing antibody response to the vaccine was measured by PRNT assay at Day 28, Day 56 and Day 90.

### Transcriptomic analysis

RNA was isolated using RNEasy micro-kit (QIAGEN) from PBMC samples taken preimmunization, and the quality of the RNA was confirmed using BGI’s protocols.

Paired-end total RNA sequencing was performed at Beijing Genomics Institute (BGI) using an Illumina NextSeq 2500 for 30 million 100 bp reads. Raw FASTQ files were processed at Case Western Reserve University with the Sekaly lab pipeline: After sequencing, reads are processed to remove Illumina adapters and low quality 3’-end bases using the Trimmomatic software (1), and then mapped to the reference mouse genome version GRCh38 using the RNA-seq optimized software STAR (2). RSeQC was then used to asses strand-specificity of reads for all transcripts (3). Transcript abundance was then estimated from unique mapped reads into raw counts using HTSeq (4), R package DESeq2 (version 1.6.2) (5)was then used to normalize read counts among samples and to identify differentially expressed genes between biological samples. A Wald test was used to evaluate the statistical relevance of the observed variations given its reproducibility between biological replicates, and a Benjamini-Hochberg correction for large number of measurements was applied to obtain adjusted p-values. Genes of interest were selected based statistical significance (p-adj > 0.05), and Bayesian shrinkage estimation was applied to the fold change to estimate effect size more accurately.

Hierarchical clustering with complete linkage was performed using Euclidean distance, and displayed using the pheatmap R package.

Preranked Gene Set Enrichment Analysis (GSEA) was performed for each contrast and or correlation against genesets extracted from the MSigDB (BROAD Institute), and Interferome databases. The shrunken fold change was used as a ranking metric Genesets found to be significantly enriched associated with the breadth of the response were considered as differentially activated pathways.

GSVA (Gene Set Variation Analysis) R package. was then used to compute a sample-level geneset enrichment z-score for genesets found to be signifcantly enriched using GSEA, on the basis of the expression level of the leading edges genes. Sample level z-scores were then used for correlation wwith other OMICs.

### Proteomic preprocessing

MRM Mass spectrometry assays was performed on a total of 228 selected plasma markers associated with inflammation and metabolism.

### Proteomic pathway analysis

Genesets were generated by the conversion of MSigDB and CHEA genesets into associated UNIPROT identifiers found in the 228 plasma proteins measured by MRM Proteomics approach. GSVA was used for each c2, c5, Hallmark and CHEA genesets on the normalized expression values. Sample level pathway scores were then compared across Breadth groups using Student’s T-test. Significant pathways were then integrated with significant selected transcriptomic pathways by using Spearman correlation on Proteomic pathway scores and Transcriptomic pathway z-scores. Significant associations were filtered using a nominal p-value <0.05.

### Metabolic profiling

Normalized expression values from Metabolon and Caprion platforms were compared across Breadth groups using a Student’s T-test. MSEA was used as an adaptation of GSEA on metabolite data by preranking the individual metabolites by their sign(t-value) * -log(p-value), and. compared to metabolic sets generated from the in-house Metabolon classifications or SMPDB (Small Molecule Pathway Database) pathways. EnrichmentMap was then used to reduce pathway redundancy of enriched pathways by generating modules of genesets on the basis of a Jaccard distance >0.25. Sample level z scores for each pathway were generated on the leading edge genes of pathways found significant in GSEA using GSVA. Those z-scores were then integrated with transcriptomic genesets selected z-scores to infer association across OMICs using Spearman correlation.

### Unsupervised Flow cytometry analysis

Individual FCS files generated by the BD FACS Diva software were imported into FlowJo Software for pregating on CD3-CD19-CD56-live cell cells. Selected events were then exported: from which an identical number of events per patient were then randomly subsampled into R. For bioinformatic analysis of flow cytometry data, a custom script was made using UMAP for RPhenograph analysis was used to to generate clusters of cells based upon their marker expression, while clusters were projected on a tSNE dimension reduction of the data, allowing for visualization of high dimensional data in two dimensions and for events to be clustered based on similar expression of flow cytometry markers which led to identification of novel cell subsets.

Frequency of events per samples was then computed per cluster and compared across Breadth groups using a Welch T-test. Median Fluorescence Intensity for each marker was computed for each cluster for comparison of expression profiles.

### Single-cell transcriptomic analysis

Publicly available datasets of 10X healthy PBMC datasets were downloaded from a prevaccination influenza cohort (https://doi.org/10.35092/yhjc.c.4753772). The top 2000 most variable genes were used as anchors to first batch correct whole PBMC datasets. Principal Component Analysis was then performed on integrated normalized data. The optimal number of components was then selected using the Elbow method, and used as input for UMAP dimension reduction and Phenograph clustering. SingleR was then used on integrated log-transformed values to infer cell type when compared to the Monaco dataset on each cluster/ SingleR was then used on individual monocyte cells to identify cell subsets. MAST software was used on raw counts to perform differential expression across groups of influenza vaccine response.

### Integrative analysis

We leveraged a projection-based approach from the R package mixOmics characterize correlations between OMICs (RNA-Seq, metabolome, proteomics, FCM). A sparse least square regression (sPLS) was used across OMICs as pairwise comparisons: a pairwise projection on the same scale allowed to quantify the Pearson correlation coefficient between the features of the two data types was calculated. To assess the probability of obtaining a Pearson correlation equal to or greater than the one observed, we derived a p value based on the distribution of the Pearson correlations calculated between all pair of features of the two data types (i.e., the statistical universe). Pearson correlations corresponding to a p value cutoff of 0.05 were considered significant.

## Data Availability

Data is not yet public, will be made public shortly with associated accession numbers in the manuscript.

## FIGURE LEGENDS

**Supplemental Figure 1.**
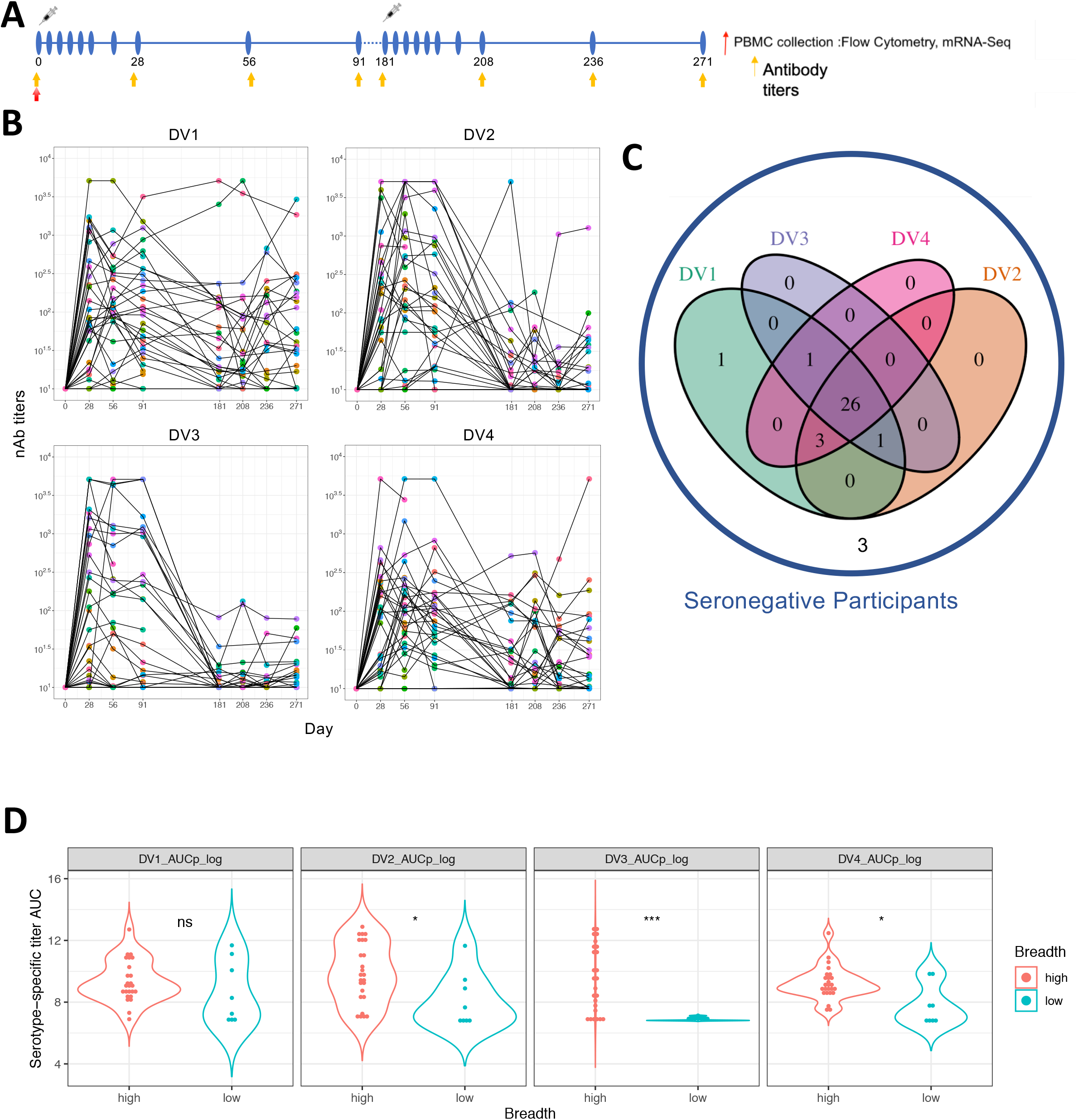
(A) Design of Stage II TV-003 Brazil Clinical Trial in Naive participants, with one primary injection at Day 0, and a second dose at Day 180. PBMCs were collected at Day 0, Day6, 15, and plasma at Day 0, (B) Neutralizing Ab responses (PRNT Assay) against each Dengue Virus serotype across Naive participants following vaccination. (C) Venn Diagram representing the number of participants that had a positive (PRNT >10) for at least one timepoint between Day 0 and Day 91 post-vaccination, against each serotype. (D) Violin Plots representing serotype-specific AUC responses per breadth group. Parametric Welch’s T-test was used to compared groups. P * < 0.05; ** < 0.01, *** <0.005

**Supplemental Figure 2. (A) Enrichment Map Interferon module leading Edge Gene network.** Edge annotation was generated using GeneMania for genes belonging to the Type I IFN modules (gray line). Tonic sensitivity classification of genes is depicted as node color, and antiviral function as a border as shown the the legend.

**Supplementary Figure 3.**
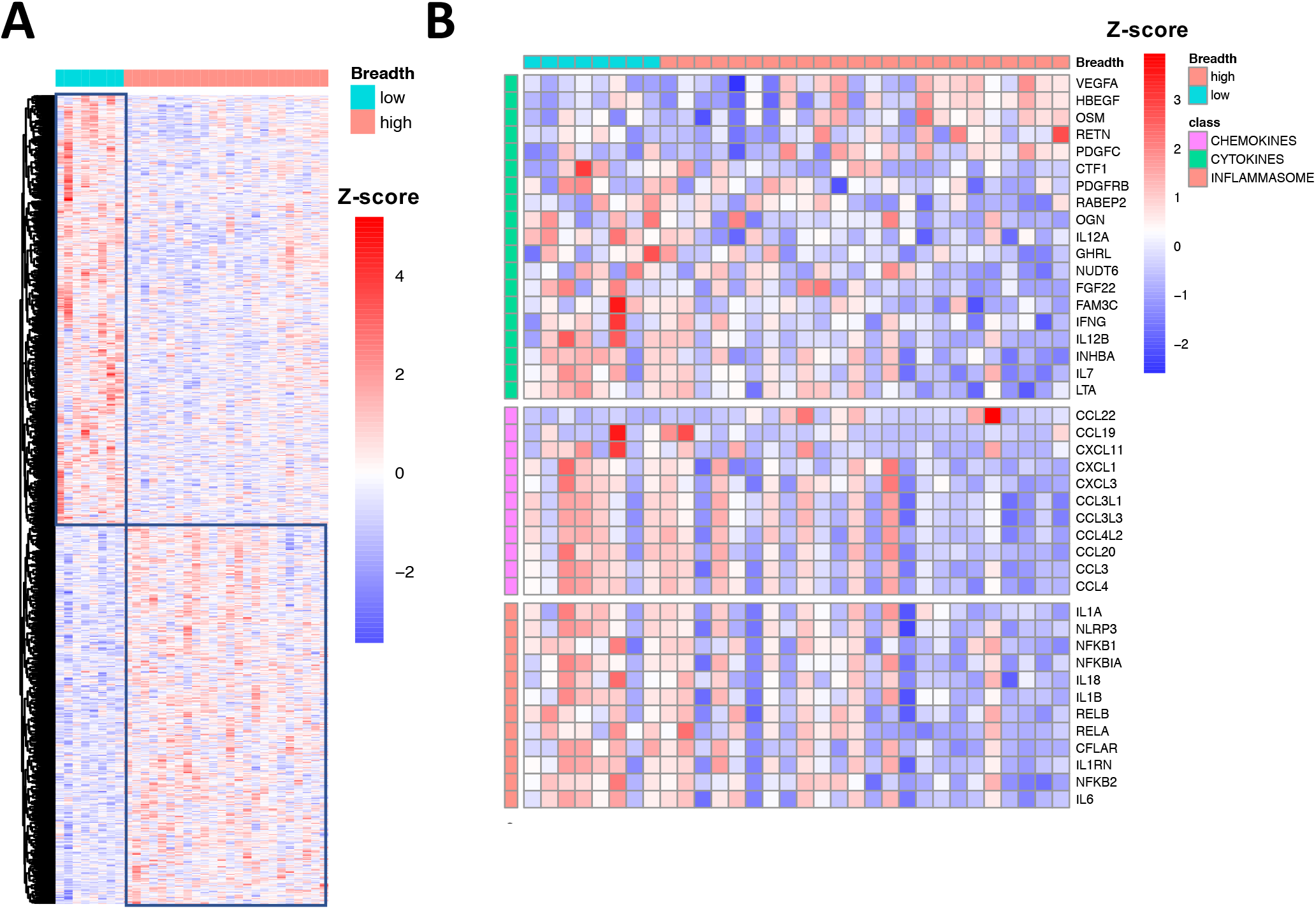
Prevaccination profilling of plasma metabolites reveals metabolic pathway activity is associated with inflammatory status b.

**Supplementary Figure 4.**
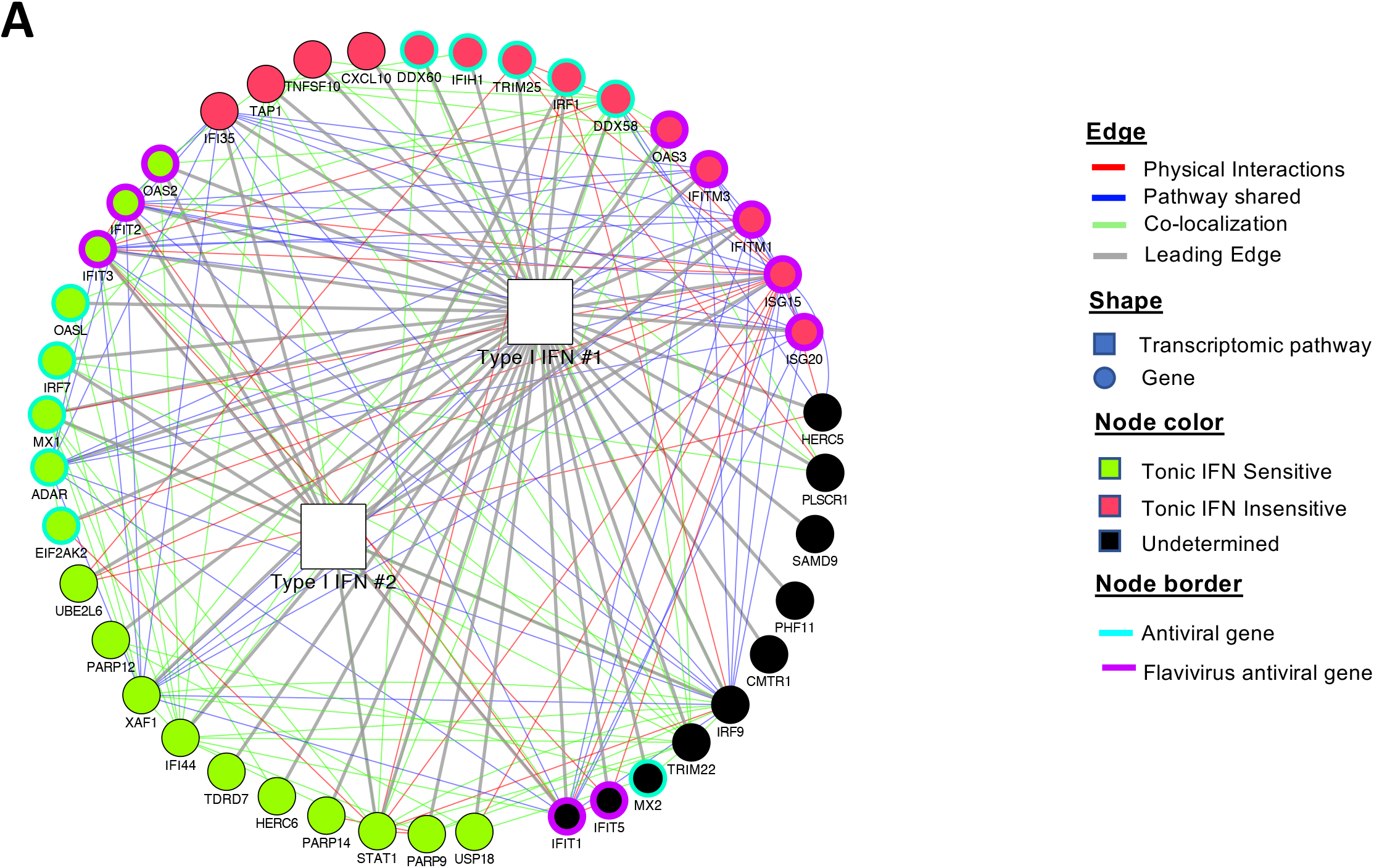
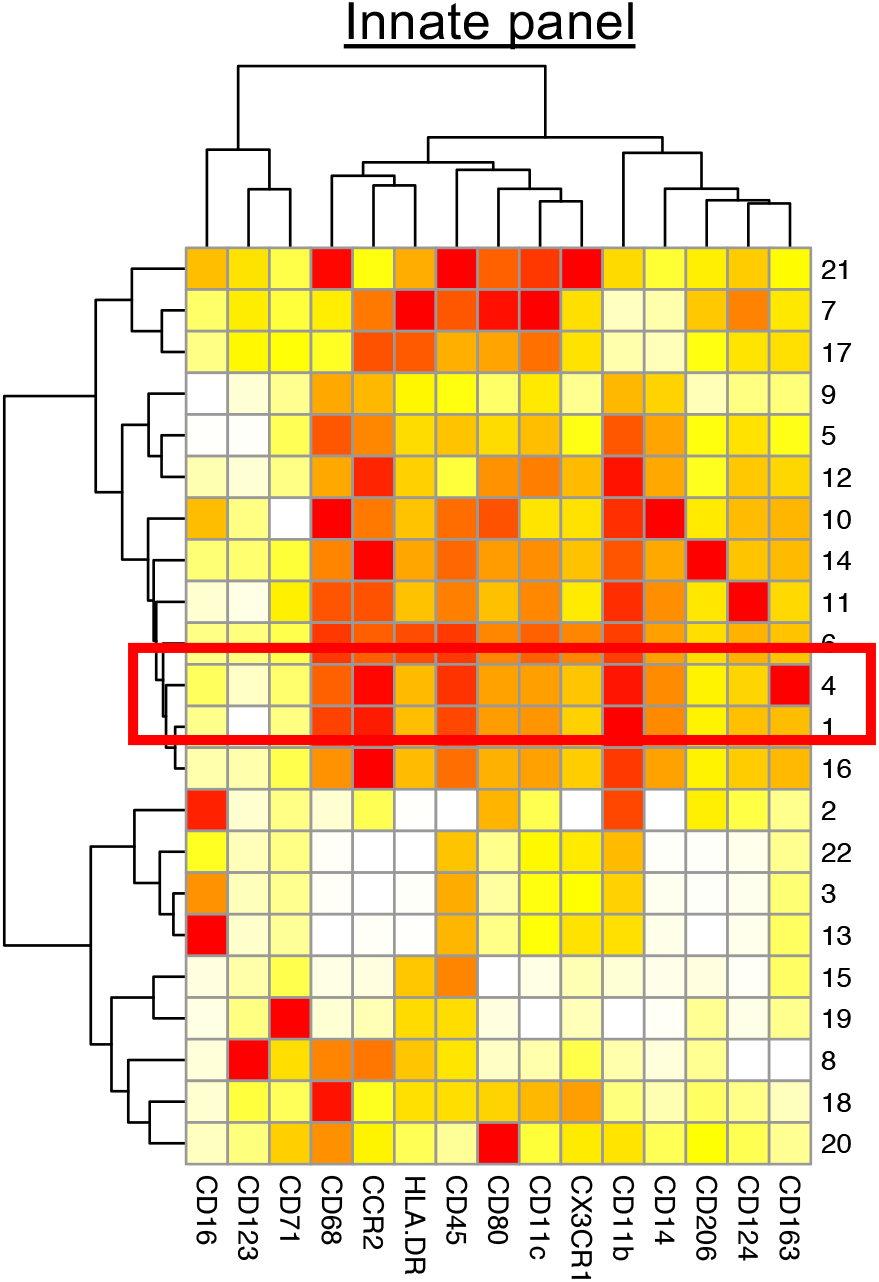
Heatmap showing scaled median fluorescence cluster profile for each marker: each row is a cluster and each column is a marker in the panel. White denotes low expression, while red denotes high expression.

**Supplementary Figure 5.**
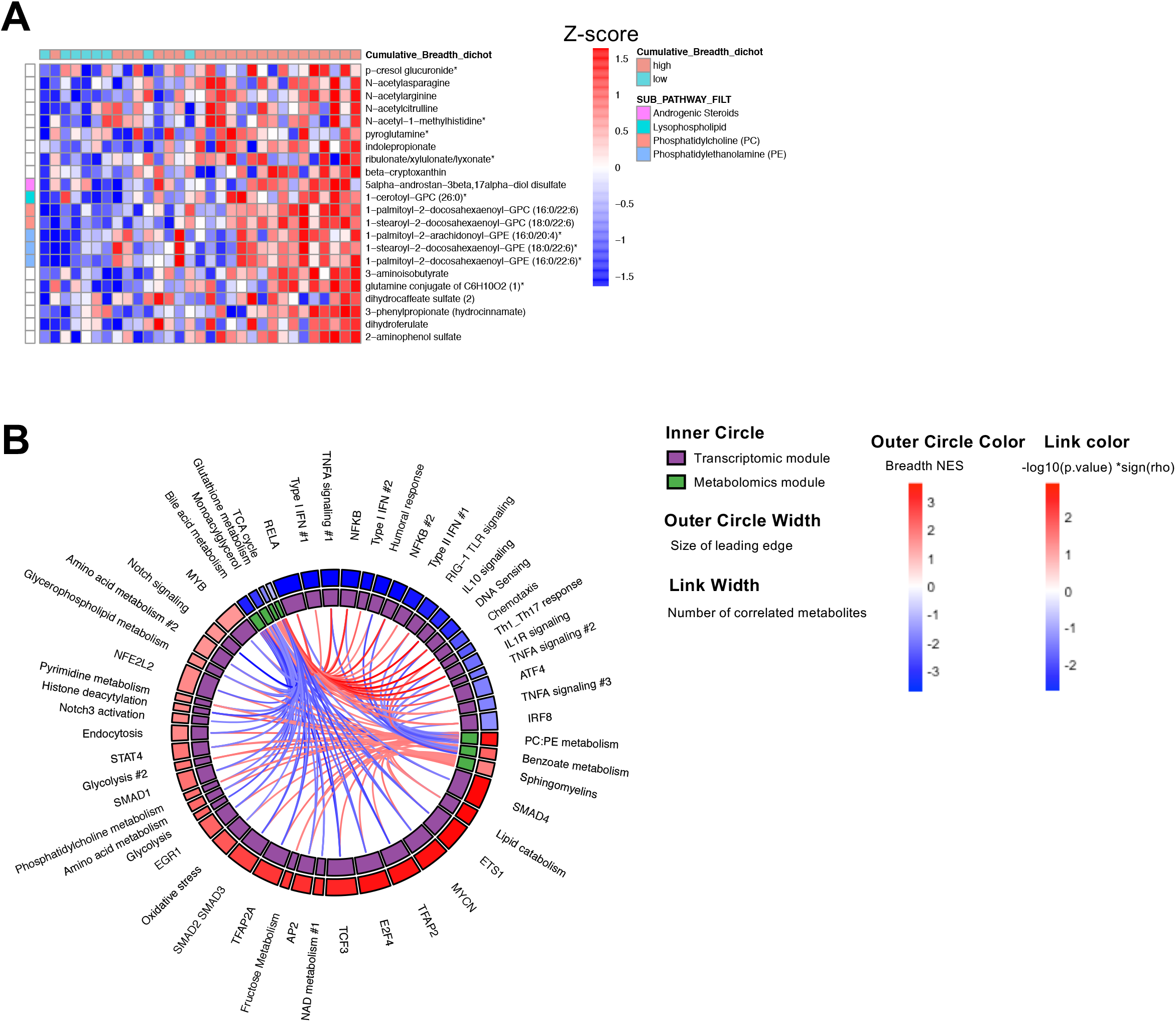
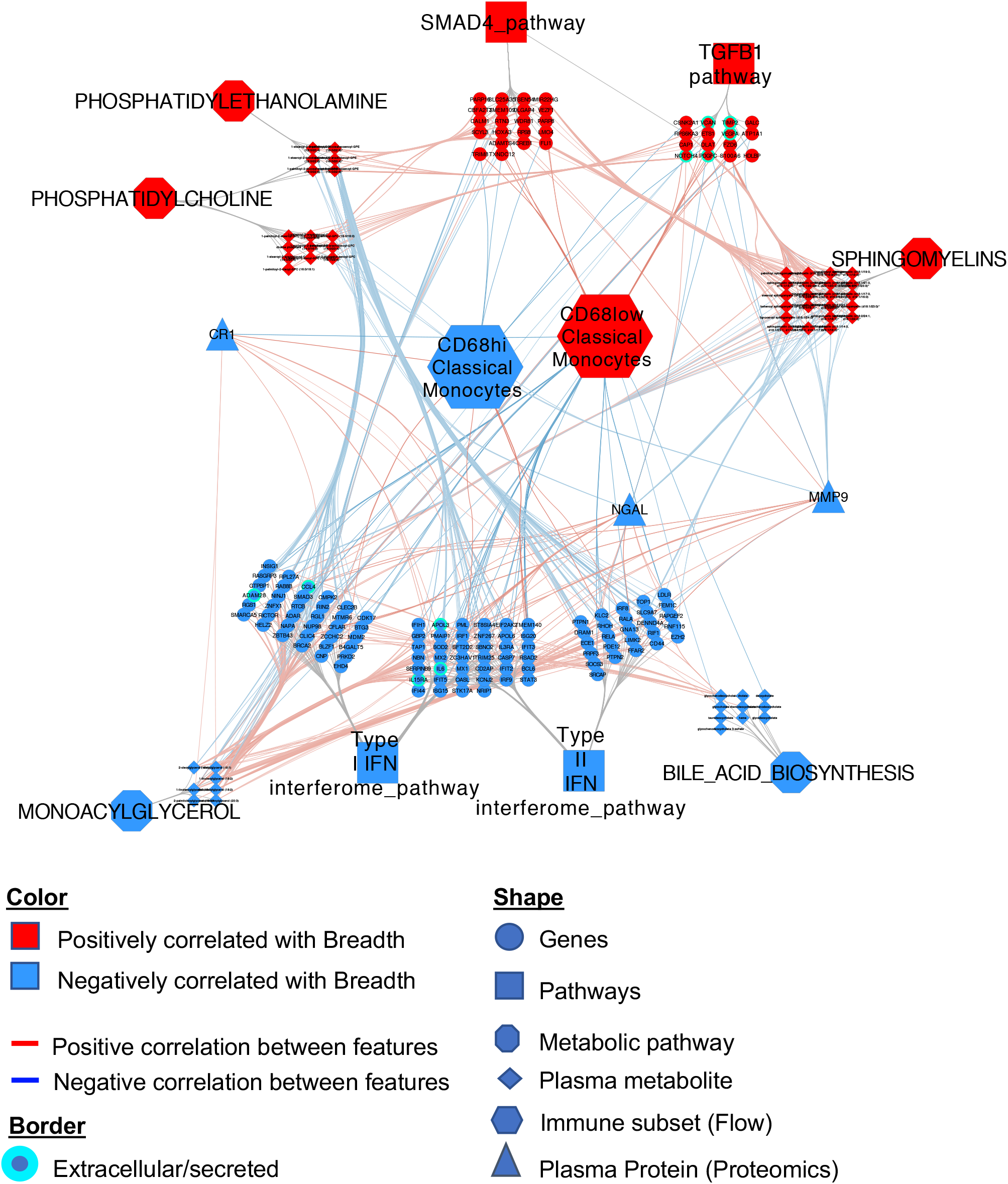

**Supplementary Figure 6.**
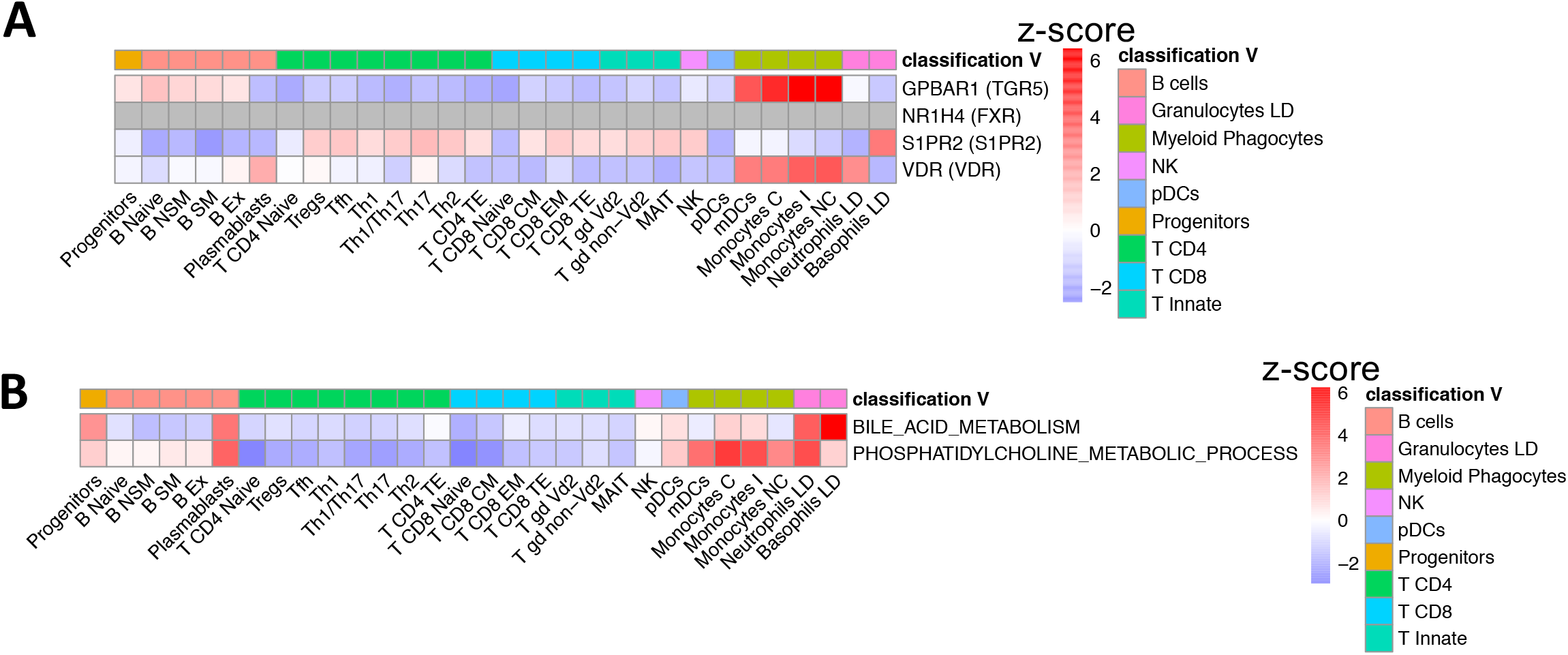

**Supplementary Table 1.** Example calculations for the Breadth outcome: the sum of serotypes with a detectable nAb titer >10 before or at Day 91 per patient (0-4) is the dichotomized into high (4) and low (<4).

| Participants | Serotype-specific nAb titers<br>(highest value across time) |  |  |  | Sum | Breadth<br>(high = 4;<br>low < 4) |
| --- | --- | --- | --- | --- | --- | --- |
|  | DV1 | DV2 | DV3 | DV4 | # serotypes:<br>titer > 10 |  |
| #1 | 1000 | 1000 | 1000 | 1000 | 4 | high |
| #2 | 100 | 100 | 100 | 100 | 4 | high |
| #3 | 1000 | 1000 | 100 | <10 | 3 | low |
| #4 | <10 | <10 | <10 | <10 | 0 | low |

**Supplementary Table 2.** Distribution of serotype-specific maximum across responder participants per timepoint. The timepoint at which each participant peaked (local maximum) per serotype was determined and summarized.

| serotype | Day 28 Max. | Day 56 Max. | Day 91 Max. | n |
| --- | --- | --- | --- | --- |
| DV1 | 15 (48.4%) | 9 (29%) | 7 (22.6%) | 31 |
| DV2 | 8 (25.8%) | 14 (45.2%) | 7 (22.6%) | 31 |
| DV3 | 17 (54.8%) | 4 (12.9%) | 6 (19.4%) | 31 |
| DV4 | 8 (25.8%) | 15 (48.4%) | 6 (19.4%) | 31 |

**Supplementary Table 3.** Distribution of the number of distinct titer maxima timepoints across serotypes.

| Number of maxima<br>timepoints per participant | n (relative proportion) |
| --- | --- |
| 1 | 4 (12.1%) |
| 2 | 18 (54.5%) |
| 3 | 11 (33.3%) |

